# Multi-pathogen serosurveillance of vaccine preventable and neglected tropical diseases in Samoa 2018-2024 to inform public health action

**DOI:** 10.64898/2026.07.20.26358474

**Authors:** Selina Rachael Ward, Harriet Lawford, Benn Sartorius, Helen Mayfield, Filipina A.L Sam, Sarah Sheridan, Robert Thomsen, Satupaitea Viali, Stefanie Vaccher, Leanne J. Robinson, Fiona Angrisano, Colleen L. Lau

**Affiliations:** Frazer Institute, The University of Queensland, Brisbane, Australia; National University of Samoa, Apia, Samoa; National Centre for Immunisation Research and Surveillance, Sydney, Australia; Samoa Ministry of Health, Apia, Samoa; Oceania University of Medicine, Apia, Samoa; Burnet Institute, Melbourne, Australia

## Abstract

**Background:** Serosurveillance can estimate the prevalence of antibodies (Ab) acquired through vaccination or current and/or past infection. Multi-pathogen serosurveillance that measures multiple Ab simultaneously, can enable identification of vulnerable sub-populations with immunity gaps to vaccine preventable diseases (VPD) and concurrent burden of neglected tropical disease (NTD), including those nearing elimination (lymphatic filariasis [LF], trachoma) and eradication (yaws). This study aimed to estimate seroprevalence and identify temporal trends of selected VPDs and NTDs in Samoa to inform targeted public health action.

**Methodology/Principal Findings:** Dried blood spots were collected from four repeated community-based surveys in eight primary sampling units (PSU) in Samoa in 2018, 2019, 2023 and 2024. Multiplex bead assays were used to detect Abs against antigens (Ag) for diphtheria, measles, rubella, tetanus, LF [*Wb123* or *Bm14*], yaws [both *Rp17* and *TmpA;* <14 years only], and trachoma *[Pgp3;* <14 years only]. Seroprevalence estimates were adjusted for sampling design and standardised for age and sex. Overall, 2,871 participants were included in this analysis. Seroprevalence of measles increased from 42% in 2018 to 95% in 2024, whereas diphtheria decreased from 79% in 2018 to 65% in 2024. Seroprevalence to yaws remained <1% for all years, whereas trachoma decreased from 21% to 7% (2018–2024). LF seroprevalence decreased between 2018 and 2024 for *Bm14* (37% to 9%) and increased for *Wb123* (10% to 22%). This study identified 15 (0.5%) individuals who were seronegative to all VPDs (7 in 2018; 8 in 2019); of these, five were seronegative to all VPDs and seropositive to at least one NTD.

**Conclusions/Significance:** Identification of sub-populations with concurrent seronegativity to VPDs and seropositivity to NTDs underscores the potential role of multi-pathogen serosurveillance in directing public health interventions to those at greatest risk. Examination of temporal patterns offer a valuable tool for measuring intervention impacts and progress towards elimination goals.

**Author Summary:** Multi-pathogen serosurveillance that measures immune responses to multiple pathogens simultaneously, can enable the identification of vulnerable sub-populations with immunity gaps to vaccine preventable diseases and concurrent burden of neglected tropical disease, including those nearing elimination (lymphatic filariasis, trachoma) and eradication (yaws). In this study, we examined 2,871 samples collected from four repeated community-based surveys in Samoa in 2018, 2019, 2023 and 2024. We observed strong gains in immunity to measles following national vaccination efforts, while immunity to diphtheria declined over the same period. For yaws and trachoma, we saw very low or decreasing patterns, consistent with progress toward elimination. Patterns for lymphatic filariasis varied depending on the antibody marker used. Additionally, this study identified a small number of people with no immunity to vaccine preventable diseases and concurrent burden with at least one neglected tropical disease, underscoring the potential role of multi-pathogen serosurveillance in directing public health interventions to those at greatest risk. Examination of temporal patterns offer a valuable tool for measuring intervention impacts and progress towards elimination goals.

## Introduction

Integrated serosurveillance (ISS), is defined by the World Health Organization (WHO) as the implementation of population-based surveys to collect and analyse samples for the simultaneous estimation of seroprevalence to multiple pathogen-specific antibodies (Ab) acquired through vaccination or current and/or past infection (1, 2). Compared with siloed serosurveillance approaches and traditional disease-specific surveillance methods, multi-pathogen serosurveillance can enable the identification of vulnerable sub-populations with immunity gaps to vaccine preventable disease (VPD) and concurrent burden of neglected tropical diseases (NTD) (3, 4), while reducing patient burden.

One approach for simultaneously measuring antibodies to multiple pathogens is the use of multiplex bead assays (MBA) that utilise florescence-based detection of Abs to up to 100 different pathogen-specific antigens (Ag) simultaneously (3, 5). The use of MBAs for ISS is gaining traction as a cost-effective tool (6, 7) to compliment traditional disease-specific surveillance method and may be a particularly beneficial approach in low-resource settings where clinical reporting and/or laboratory facilities may be limited (8–10). Furthermore, this approach allows for the opportunity to detect circulating infections that are not routinely monitored, particularly those targeted for elimination or eradication (11). For VPDs, serosurveillance can provide more accurate estimates of population immunity compared to relying on vaccination coverage based on historical records (12). Combining seroprevalence estimates with epidemiological and geographic data enables multiple public health applications, including simultaneous identification of infection hotspots, characterisation of pockets of immunity gaps or waning immunity to VPDs (13), measuring the impact of interventions (14, 15), and guiding disease elimination efforts (16).

In 2003, the WHO Western Pacific Region (WPR) set the goal to eliminate measles in the region by 2012, urging all member states to achieve and maintain 95% population immunity in each birth cohort (17). Similarly, the goal to eliminate rubella was declared in 2014 following an estimation of approximately 9000 congenital rubella cases within the region (18). Samoa, a lower-middle income tropical island nation in the WPR (19), operates within a healthcare system that includes public, private and traditional sectors. However, the distribution of services is highly uneven with limited focus on primary health care of care (20, 21). Similar to many other Pacific Islands and Territories (PICT) Constrained by limited laboratory capacity, traditional disease surveillance systems in Samoa have largely operated in silos with a large emphasis on syndromic case reporting within the Pacific Public Health Surveillance Network (22). While most healthcare challenges relate to noncommunicable diseases (23), Samoa continues to face infectious disease outbreaks and large variation in reported immunisation coverage for VPDs.

Coverage of routine childhood vaccinations in Samoa, including measles, mumps and rubella (MMR) and diphtheria, tetanus and pertussis (DTP) have been variable over the past decade. Following two vaccine-associated paediatric deaths in July 2018, community mistrust in the safety of vaccination saw rates had fall to 40% for first-dose measles containing vaccination (MCV) and 28% for second-dose (24). Following the measles outbreak in 2019, Samoa initiated a catch-up vaccination campaign targeting individuals aged 6-50 years, with a reported 187,369 MCV doses given, achieving a coverage of 93.3% (25).

Lymphatic filariasis (LF) remains endemic in Samoa despite ongoing elimination efforts led by the Ministry of Health (MoH), with support from the Pacific Programme to Eliminate Lymphatic Filariasis (PacELF) program between 1999-2007 (26, 27). Prior to intense control efforts in the 1950s-1960s, Samoa was also considered endemic for trachoma and yaws. Recently, a retrospective review of ophthalmic patient records between 2011-2013 found no documented cases of trachoma (28), however, the current status is unknown. Importantly, following disruption to routine health services, including NTD elimination programs during the COVID-19 pandemic (29), understanding population immunity to both VPDs and NTDs is critical for identifying residual vulnerabilities and directing targeted public health interventions to achieve elimination efforts.

The Surveillance and Monitoring to Eliminate Lymphatic Filariasis and Scabies in Samoa (SaMELFS) project is an operational research program designed to monitor and evaluate the effectiveness of nationwide triple-drug mass drug administration (MDA) on reducing LF prevalence in Samoa (26, 27, 30). Multiple surveys have been conducted since 2018 (31, 32). Previous seroepidemiology studies using data obtained through these surveys have provided important insights into the national prevalence of NTDs and VPDs in 2018 (33), the spatial distribution and clustering of VPDs in 2018 and 2019 (13), and comparison between serological and vector indictors of LF transmission between 2018-2024 (34). Here, we aimed to build upon this work and examine the multi-year seroprevalence of selected VPDs and NTDs within eight primary sampling units (PSU). Collating MBA data from 2018, 2019, 2023 and 2024, our specific objectives were to: i) estimate the annual seroprevalence and examine temporal trends for VPDs (measles, rubella, tetanus, diphtheria) and NTDs (LF, yaws, trachoma); ii) examine seropositivity to combinations of VPD and NTDs; and ii) identify sex and age-group specific differences in seroprevalence.

## Methods

### Ethics statement

Ethics approvals for years 2018-2023 were granted by the Samoa Ministry of Health (MOH) and The Australian National University Human Research Ethics Committee (2018/341) and ratified by The University of Queensland (2021/HE000895). Ethics approvals for 2024 were granted by the Samoa MOH and The University of Queensland (2024/HE001263). The studies were conducted in close collaboration with the Samoa MOH, the WHO country office in Samoa, and the Samoa Red Cross. Written informed consent was obtained from adult participants, and verbal assent was obtained from participants aged <18 years with formal written consent obtained from a parent or guardian. All research activities were performed in accordance with the relevant guidelines and regulations.

### Study setting

With a population of 218,019 in 2024 (19), Samoa is comprised of two main islands with over 330 villages, contained within 11 political districts (35). The Samoan health care system is centrally overseen and organised by the MOH, delivered through the national referral hospital Tupua Tamasese Meaole (TTM) Hospital, and a network of distract hospitals, community health centres and private medical clinics. Alongside this, several non-governmental organisations and religious organisations are engaged in healthcare delivery and advocacy (36).

### Data source

In 2018 and 2019, the SaMELFS surveys were designed to be nationally representative, containing 35 PSUs and have been described in detail elsewhere (26, 32, 37, 38). Surveys were not conducted in 2020-2022 due to the COVID-19 pandemic. In 2023 and 2024, smaller, targeted surveys were conducted in eight purposively-selected PSUs (to represent zero, low, moderate, and high LF prevalence) that had previously been sampled in 2018 and 2019 (31). In all years, participant demographic information was collected using a standardised electronic questionnaire. Eligibility criteria included: Age >5 years, lived in Samoa for a total of 12 months over the past 5 years, and current resident of the household. All eligible participants had a finger prick sample of up to 400µL of blood collected into heparin microtainers to be used to conduct point-of-case testing for filarial antigen (Alere™ Filariasis Test Strips in 2018 and 2019 and Standard Q Filariasis Antigen Test [QFAT] in 2023 and 2024) and to prepare dried blood spots (DBS) by pipetting 10µL of blood onto each ear (up to six ears total) of a Cellabs Tropbio Filter Paper Blood Collection Disk™. The design, components and participants recruited in each year are described in further detail in Supplementary Material Text 1.

### Laboratory analysis

To allow comparison across multiple years, antigens used to examine Abs included in this analysis (Table 1) were restricted to: LF (*Bm14* and *Wb123*), trachoma (*Pgp3*), yaws (*Rp17* and *TmpA*), tetanus (*Tet*), diphtheria (*Dip*), measles (*MeV*), rubella (*RuV*). Samples collected in 2018 and 2019 were analysed at the US Centers for Disease Control and Prevention (US CDC) Division of Parasitic Diseases and Malaria, Atlanta, USA. Samples collected in 2023 and 2024 were analysed at the Vector-Borne Diseases and Tropical Public Health laboratory at the Burnet Institute, Melbourne, Australia. In brief, DBS were eluted into 96-well plates, diluted to a final concentration of 1:400 and assay sample plates were read on a Bio-Plex 200 MBA instrument (Bio-Rad, Hercules, CA).

**Table 1:** Antigen characteristic list for pathogens examined using multiplex bead assays for samples collected in the Surveillance and Monitoring to Eliminate Lymphatic Filariasis and Scabies in Samoa (SaMELFS) studies 2018-2024.

| *Disease* | *Pathogen* | *Antigen/s* | *Description* | *References* |
| --- | --- | --- | --- | --- |
| *Lymphatic Filariasis* | *Brugia malayi* | SXP-1 (*Bm14*) | Marker of historical or current infection. *Bm14* Ab is proposed to reflect recent exposure to infective larvae and the presence of adult worms and is suggested to be better suited to detect recrudescence or persisting transmission of *W. bancrofti* after MDA cessation. | (39-43) |
|  | *Wuchereria bancrofti* | Larval antigen (*Wb123*) | Marker of historical or current infection. It has been proposed that the response to *Wb123* is developed after repeated larval stimulation, possibly indicating ongoing transmission or prepatent infection. There is some evidence that decrease in *Wb123* antibody levels after treatment occurs quicker than *Bm14*, and is inversely related to the infection load before treatment. | (42, 44-49) |
| *Trachoma* | *Chlamydia trachomatis* | pCT03 ORF (*pgp3*) | Marker of historical or current infection. Seroprevalence in older age groups may represent sexually transmitted infection (urogenital chlamydia), therefore analysis was restricted to <14 years. Previously, seropositivity was determined if positive to both *pgp3* and *CT694* (T3SS substrate), however recent evidence suggests that positivity to *pgp3* Ab alone is sufficient. There is evidence that *pgp3* Ab seropositivity can remain stable for over one year in low-endemicity settings, however a single childhood infection alone may not be sufficient for seropositivity. | (50-54) |
| *Yaws* | *Treponema pallidum pertenue* | Yaws (*Rp17*) | Marker of historical infection to *T. pallidum subsp pallidum* (syphilis) or *T. pallidum subsp pertenue* (yaws). Unable to differentiate between subspecies due to cross-reactivity. Seroprevalence analysis was restricted to <14 years. | (55-58) |
|  |  | Treponemal membrane protein A (*TmpA*) | Marker of current or recent infection to *T. pallidum subsp pallidum* (syphilis) or *T. pallidum subsp pertenue* (yaws). Unable to differentiate between subspecies due to cross-reactivity. Seroprevalence analysis was restricted to <14 years. There is evidence that *TmpA* Ab tend to decrease within 6 months to a year of treatment. | (55, 57, 59-62) |
| *Measles* | Measles | Whole virus (*MeV*) | Marker of vaccination or historical infection. Cannot distinguish between vaccine-acquired immunity or natural infection. Antibody titres equivalent to | (63-66) |
| *Rubella* | Rubella | Whole virus<br>(*RuV*) | $\geq 0.01$ IU/mL shows good correlation with protection against symptomatic disease; however, cases of secondary vaccine failure may still occur. | |
| *Diphtheria* | Diphtheria | Toxoid (*Dip*) | Marker of vaccine-acquired immunity. Antibody titres equivalent to $\geq 0.01$ | (67, 68) |
| *Tetanus* | Tetanus | Toxoid (*Tet*) | IU/mL for tetanus and diphtheria are considered to provide minimum protection against symptomatic infection. |  |

### Classification of seropositivity

For each Ag, median fluorescence intensity minus background (MFI-bg) signal was used to determine seropositivity. Thresholds for seropositivity (Supplementary Material Table 1) were determined through one of four methods: (1) the mean plus three standard deviations (SD) of the MFI-bg of known negative sera; (2) receiver operating characteristics (ROC) derived from MFI-bg cut-offs based on known positive and negative controls; (3) standard curves using reference serum standards from the International Reference Materials of the World Health Organization; or (4) Finite mixture model/s.

### Statistical analysis

#### Analytical Dataset and Inclusion Criteria

Data cleaning and analysis was performed in Stata (StataCorp, Version 17.0, College Station, TX). Analysis for this current study was restricted to participants recruited through random household-based sampling within the eight purposefully selected PSUs (based on 2019 LF Ag prevalence: Zero – Vaivase Tai and Mutiatele and Saleaaumu; Low – Tuanai and Fusi; Medium – Vaiusu and Falefa, and High – Falaesiu and Lauli’i) (31). The participants included in analysis for each year are shown in Supplementary Material Text 1. To examine for sampling bias across years, we performed chi-square testing on the proportion of participants in each sex and age group by PSU.

#### Estimation of seroprevalence by year and assessment of temporal change

Weighting for probability of household selection was calculated as the number of households selected in each PSU and year, over the total number of households in the PSU according to the census (2016 census for years 2018 and 2019, and 2021 census for years 2023 and 2024). Standardisation to age and sex was calculated using the proportion of people in each age-group and sex according to the census, with weighting calculated as the inverse of this proportion (Calculations are included in Supplementary Material Table 2). Seroprevalence estimates and 95% CI were adjusted for PSU selection and household sampling using the *pweight* command, and weighting for sex and age groups were adjusted for using the *stdize* and *stdweight* postestimation standardization commands (33). Logistic regression was performed at the individual level from binomial sampling process to calculate odds ratios (OR) and 95% CI with 2018 as the reference year. Multiple–testing correction was performed using the Westfall–Young permutation method (*wyoung*) with seropositivity as the outcome and survey year as the predictor. One hundred permutation replicates were used to obtain adjusted p–values.

#### Analysis of poly–seropositivity patterns

Seropositivity profiles for individuals were determined with those seropositive to more than one VPD and/or NTD considered poly-seropositive.

#### Stratified analyses by age and sex and definitions of sex and gender

Combining seropositivity data for all years, age (5-10; 11-20; 21-30; 31-40; 41-50; 51-65; 65+ years) and sex (male and female) group differences were examined using logistic regression. When conducting fieldwork surveys, the questionnaire asked specifically “Sex – choose (male, female)”, however, the Samoan census states “Is the gender identity that (name) is identify with, the same as (his/her) sex registered at birth?”(69). Therefore, in this analysis, we have used the term “sex” to refer to “gender”.

## Results

### Study participants

Overall, 2,871 participants were eligible for inclusion (551 [19.2%] in 2018, 625 [21.7%] in 2019, 598 [20.8%] in 2023, 1,097 [38.3%] in 2024). A summary of participants and demographic characteristics are shown in Table 2. For all PSUs and households combined, there were no statistically significant changes in the age or sex of participants (Pearson chi2=27.09; p=0.168), indicating no sampling bias across years (Supplementary Material Table 3).

**Table 2:** Number of participants and demographics of participants included in analysis from the Surveillance and Monitoring to Eliminate Lymphatic Filariasis and Scabies in Samoa (SaMELFS) studies 2018-2024.

| ***Demographics*** | ***2018*** | ***2019*** | ***2023*** | ***2024*** |
| --- | --- | --- | --- | --- |
| *Total participants* | 551 | 525 | 598 | 1097 |
| ***Sex*** |  |  |  |  |
| *Male* | 284 (45.4%) | 275 (49.9%) | 267 (44.6%) | 505 (46.1%) |
| *Female* | 341 (54.5%) | 276 (50.1%) | 331 (55.3%) | 591 (53.9%) |
| ***Age group*** |  |  |  |  |
| *5-14 years* | 26 (36.1%) | 181 (32.8%) | 204 (34.1%) | 401 (36.5%) |
| *15-29 years* | 184 (29.4%) | 149 (27.0%) | 151 (25.2%) | 282 (25.7%) |
| *30-64 years* | 182 (29.1%) | 185 (33.5%) | 209 (34.9%) | 349 (31.8%) |
| *65+ years* | 33 (5.2%) | 36 (6.5%) | 34 (5.6%) | 65 (5.9%) |

### Seroprevalence by year and temporal change

All seroprevalence estimates and 95% confidence intervals (CI) are shown in Table 3. Odds of seroprevalence compared to 2018 are shown in Supplementary Material Table 4.

**Table 3:** Seroprevalence estimates ^a^ (%) and 95% confidence intervals to vaccine preventable diseases (measles, rubella, diphtheria and tetanus) and neglected tropical diseases (trachoma, yaws, and lymphatic filariasis) in Samoa 2018, 2019, 2023 and 2024. a-Adjusted to primary sampling unit (PSU) and household sampling probability and standardised for sex and age.

|  |  | *Seroprevalence (95% confidence interval)* |  |  |  |
| --- | --- | --- | --- | --- | --- |
|  | *Year* | **2018** | **2019** | **2023** | **2024** |
| ***Vaccine preventable diseases*** |  |  |  |  |  |
|  | *Measles* | 42.47 (36.14-49.05) | 46.07 (39.17-53.13) | 98.83 (95.92-99.67) | 96.60 (95.85-97.22) |
|  | *Rubella* | 73.94 (62.22-83.018) | 80.07 (72.80-85.77) | 73.27 (65.68-79.70) | 98.87 (96.89-99.59) |
|  | *Diphtheria* | 79.18 (74.18-83.42) | 78.28 (73.29-82.56) | 68.06 (62.85-72.86) | 65.06 (59.84-69.94) |
|  | *Tetanus* | 90.36 (84.22-94.28) | 87.63 (79.56-92.80) | 94.41 (91.10-96.54) | 92.33 (89.32-94.54) |
| ***Neglected tropical diseases (<14 years)*** |  |  |  |  |  |
|  | *Trachoma (pgp3)* | 21.11 (13.66-31.14) | 10.46 (8.10-13.42) | 9.04 (4.91-16.06) | 7.18 (3.91-12.82) |
|  | *Yaws (rp17)* | 0.00 (0.00-0.00) | 5.70 (1.57-18.57) | 1.06 (0.24-4.53) | 2.66 (1.17-5.93) |
|  | *Yaws (TnpA)* | 0.58 (0.08-3.89) | 1.81 (0.62-5.15) | 15.30 (10.16-22.40) | 5.17 (3.01-8.73) |
|  | *Yaws (both rp17 & TnpA)* | 0.00 (0.00-0.00) | 0.00 (0.00-0.00) | 0.13 (0.02-0.94) | 0.23 (0.08-0.72) |
| ***Lymphatic filariasis (LF)*** |  |  |  |  |  |
|  | *LF (Bm14)* | 37.98 (32.34-43.96) | 29.24 (24.44-34.55) | 14.85 (11.1-19.59) | 9.92 (7.72-12.66) |
|  | *LF (Wb123)* | 10.08 (7.09-14.15) | 10.7 (7.57-14.91) | 9.83 (6.99-13.64) | 22.93 (19.49-26.77) |
|  | *LF (either Bm14 or Wb123)* | 38.23 (32.59-44.21) | 30.42 (25.52-35.81) | 16.54 (12.63-21.36) | 25.12 (21.57-29.03) |
|  | *LF (both Bm14 and Wb123)* | 9.83 (6.86-13.89) | 9.51 (6.66-13.41) | 8.14 (5.56-11.77) | 7.73 (5.85-10.15) |

**Table 3:** Number of participants (n; %) and demographics of participants seronegative to vaccine preventable diseases (measles, rubella, diphtheria, tetanus) and seropositive to at least one neglected tropical disease (yaws, trachoma, lymphatic filariasis) in 2018 and 2019.

|  | 2018 | 2019 |
| --- | --- | --- |
| ***Seronegative to all VPDs\**** |  |  |
| ***Number (%)*** | 7 (1.3%) | 8 (1.28%) |
| ***Sex and age group (years)*** |  |  |
| *Female 5-14* | - | 4 (50.00%) |
| *Female 15-29* | 4 (57.14%) | - |
| *Female 30-64* | - | - |
| *Female 65+* | - | - |
| *Male 5-14* | - | 2 (25.00%) |
| *Male 15-29* | 2 (28.57%) | 2 (25.00%) |
| *Male 30-64* | 1 (14.29%) | - |
| *Male 65+* | - | - |
| ***Seronegative to all VPDs\* and at least one NTD^*** |  |  |
| ***Number (%)*** | 2 (0.36%) | 3 (0.48%) |
| ***Sex and age group:*** |  |  |
| *Female 5-14 years* | - | 2 (66.67%) |
| *Female 15-29* | 2 (100%) | - |
| *Male 5-14 years* | - | - |
| *Male 15-29 years* | - | 1 (33.33%) |
| * Vaccine preventable diseases (VPDs): measles, rubella, diphtheria, tetanus<br>^Neglected tropical disease (NTD): All ages - Lymphatic filariasis (LF) either Bm14 or Wb123 Ab; <14 years - yaws (either rp17 or TmpA Ab) or trachoma (Pgp3 Ab) |  |  |

#### VPDs

Seroprevalence to measles increased significantly from 42.5% in 2018 to 98.8% in 2023 and 96.6% in 2024 (OR=20.3 [reference: 2018]; 95% CI: 13.6-29.4). A similar increase was observed for rubella seroprevalence, increasing from 73.94% in 2018 to 98.9% in 2024 (OR=21.2; 95% CI: 12.4-36.2). In contrast, seroprevalence to diphtheria decreased from 79.2% in 2018 to 68.0% in 2023 and 65.0% in 2024 (OR=0.3; 95% CI: 0.2-0.5).

#### NTDs

The observed decline in seroprevalence for trachoma (*Pgp3* Ab) from 21.1% (95% CI: 13.6-31.1) in 2018 to 9.0% in 2023 and 7.1% in 2024, was not statistically significant. The increase in seroprevalence to yaws (seropositive to both *Rp17* or *TmpA* Ab) in those aged <14 years from to 0.0% in 2018 and 2019 to 0.23% (0.0-0.7) in 2024, was not statistically significant (p=0.98). LF seroprevalence to *Bm14* Ab decreased significantly from 37.9% in 2018 to 14.8% in 2023 (OR=0.2; 95% CI: 0.1-0.3) and 9.9% in 2024 (OR=0.1; 95% CI: 0.1-0.2). Seroprevalence to *Wb123* Ab increased significantly from 10.0% in 2018 to 22.9% in 2024 (OR=1.9; 95% CI: 1.4-2.6).

### Poly-seropositivity

The number of participants seropositive to combinations of VPDs (measles, rubella, diphtheria and tetanus) are shown in Figure 1. The demographics and number of participants seronegative to all VPDs and/or seropositive to at least one NTD are shown in Table 4 (additional combinations included in Supplementary Material Table 5). Across all years, 15 individuals were seronegative to all VPDs (7 in 2018 and 8 in 2019), with a median age of 16 years (IQR: 10-20 years). No individuals seronegative to all VPDs were identified in 2023 or 2024. Among the 15 individuals found to be seronegative for all four VPDs, this study identified five individuals (2 in 2018 and 3 in 2019) seronegative to all VPDs and seropositive to at least one NTD (median age=16; IQR: 14-20 years), of which, four (80.0%) were female.

**Figure 1:**
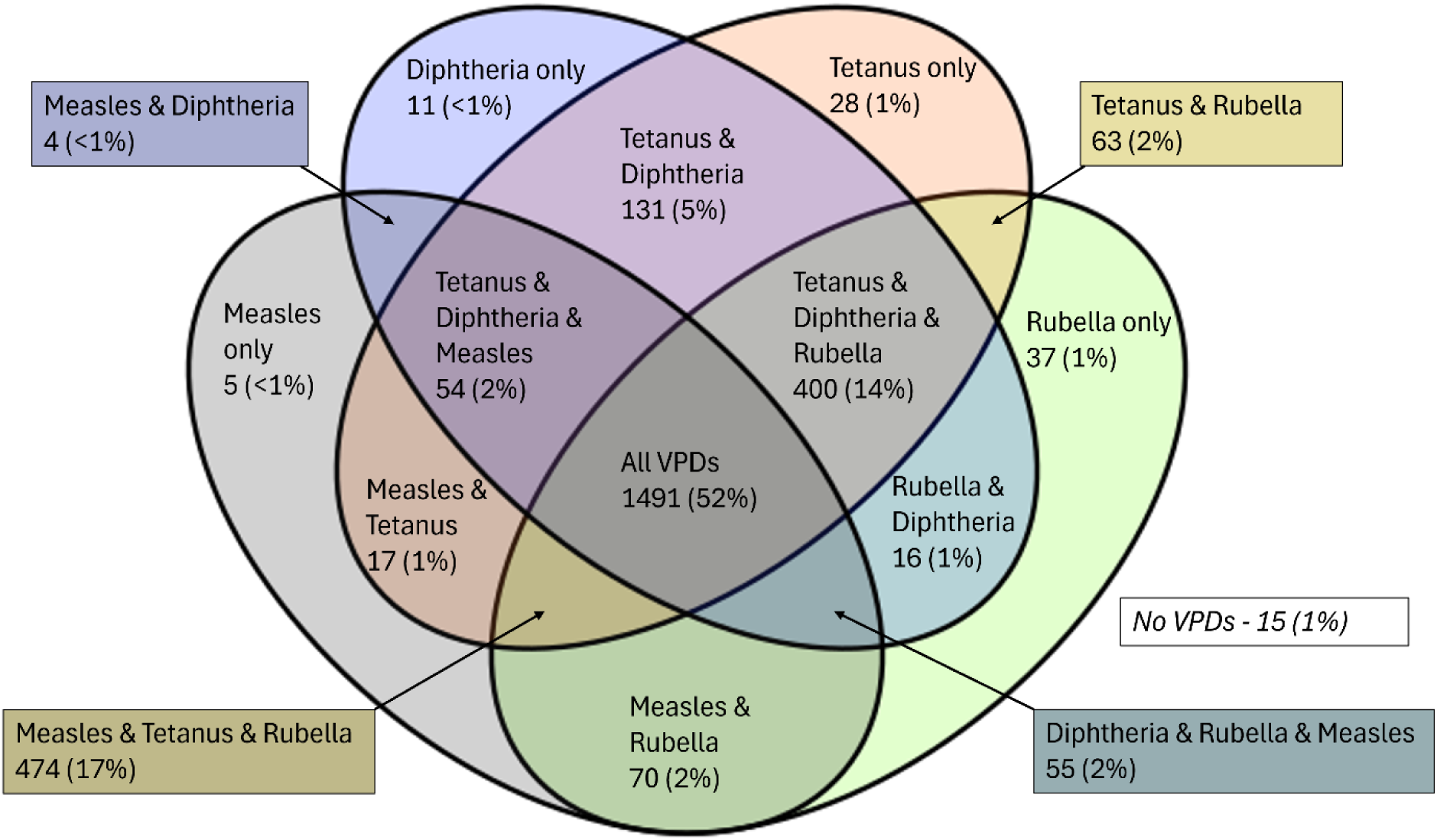
Venn diagram of number (% of total) of participants seropositive to combinations of vaccine preventable diseases (measles, rubella, diphtheria and tetanus) from the Surveillance and Monitoring to Eliminate Lymphatic Filariasis and Scabies in Samoa (SaMELFS) studies 2018-2024.

### Sex and age-group specific differences in seroprevalence

Yearly seroprevalence to VPDs by sex and age groups are shown in Figure 2 with yearly seroprevalence to LF by sex and age groups shown in Figure 3. For all years combined, the odds of seropositivity compared to the 5-10 years age group for VPDs and LF are shown in Figure 4.

**Figure 2:**
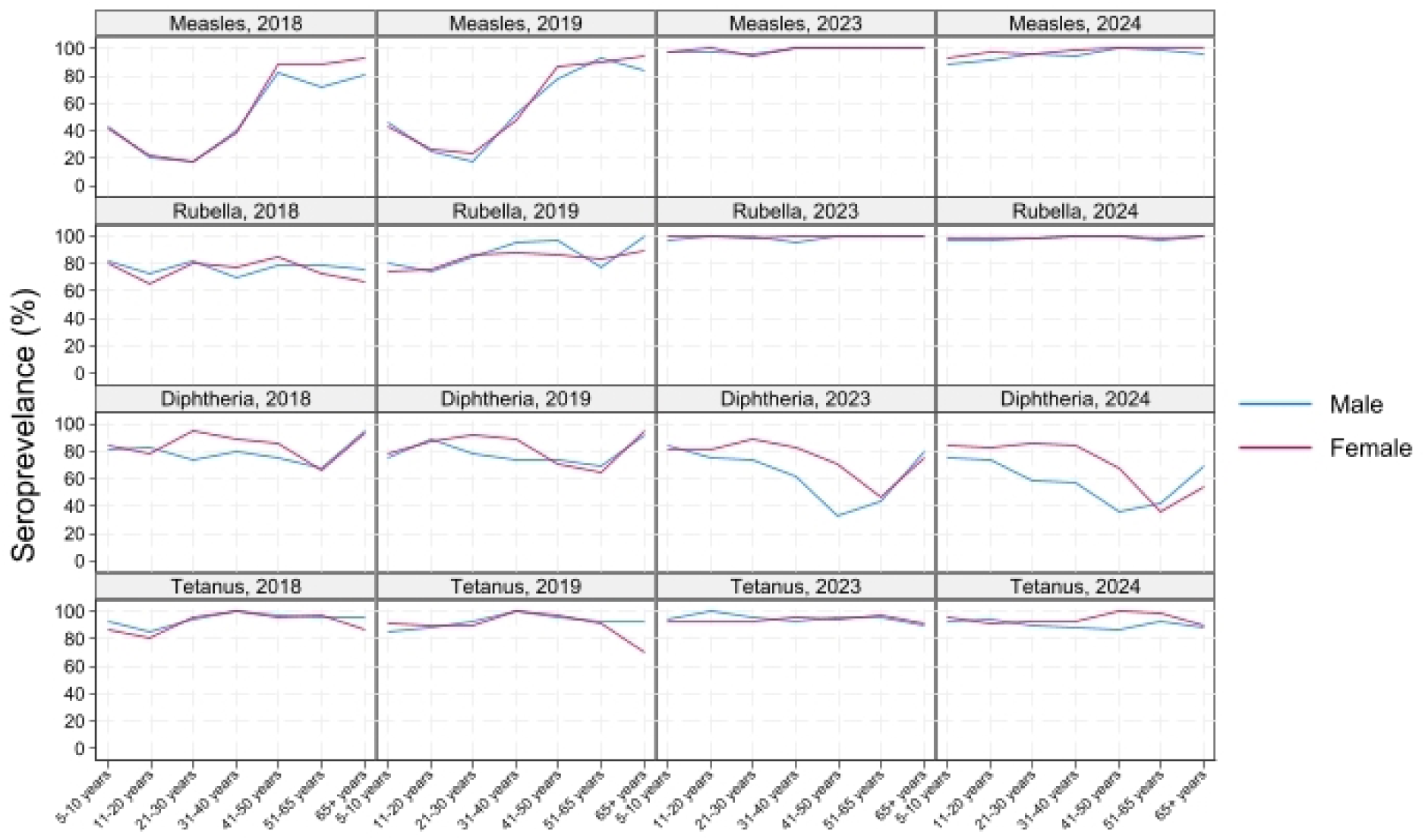
Seroprevalence (%) to vaccine preventable diseases (measles, rubella, diphtheria, tetanus) by age group and sex in Samoa for years 2018, 2019, 2023 and 2024.

**Figure 3:**
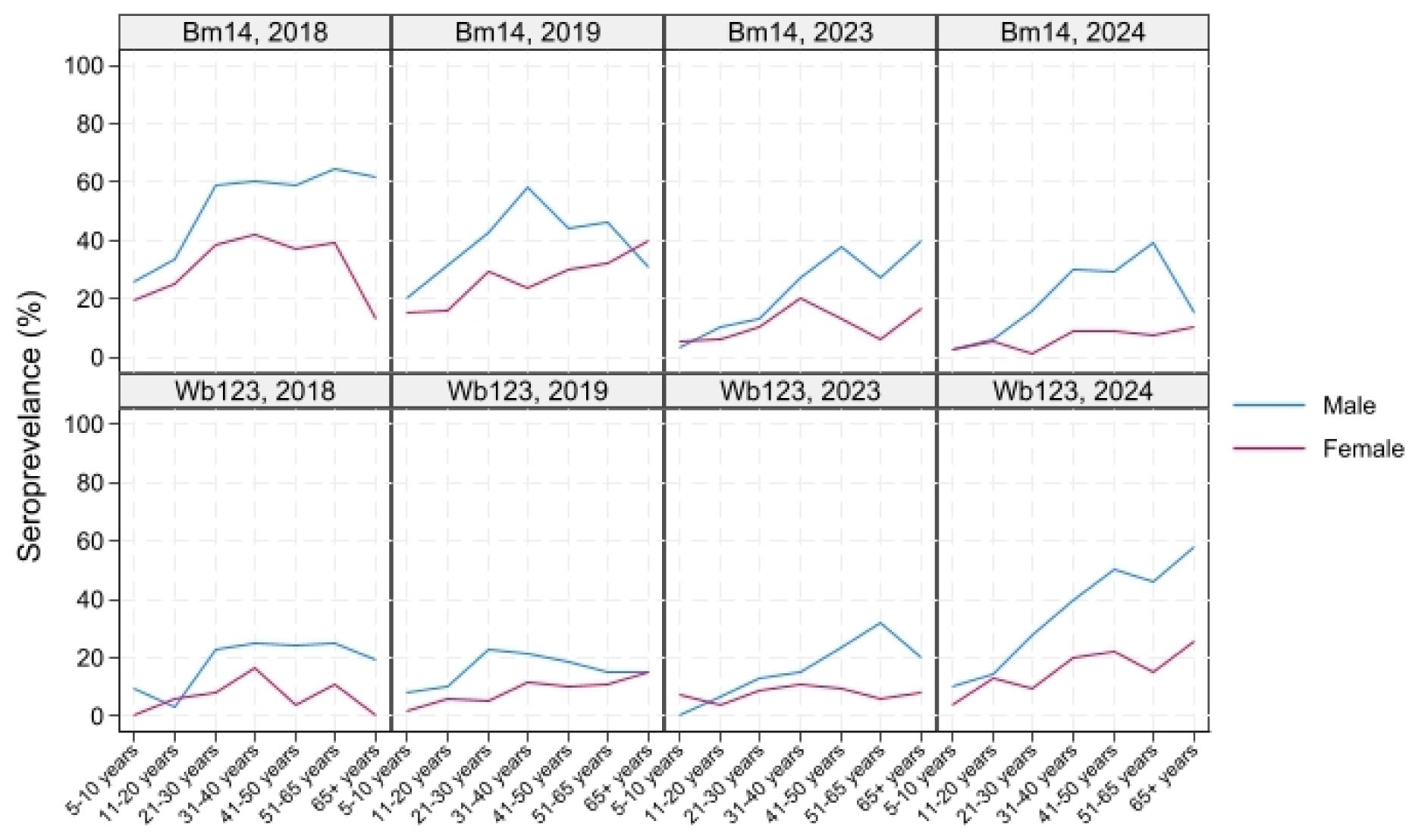
Seroprevalence (%) to lymphatic filariasis antibodies (Bm14, Wb123) by age group and sex in Samoa for years 2018, 2019, 2023 and 2024.

**Figure 4:**
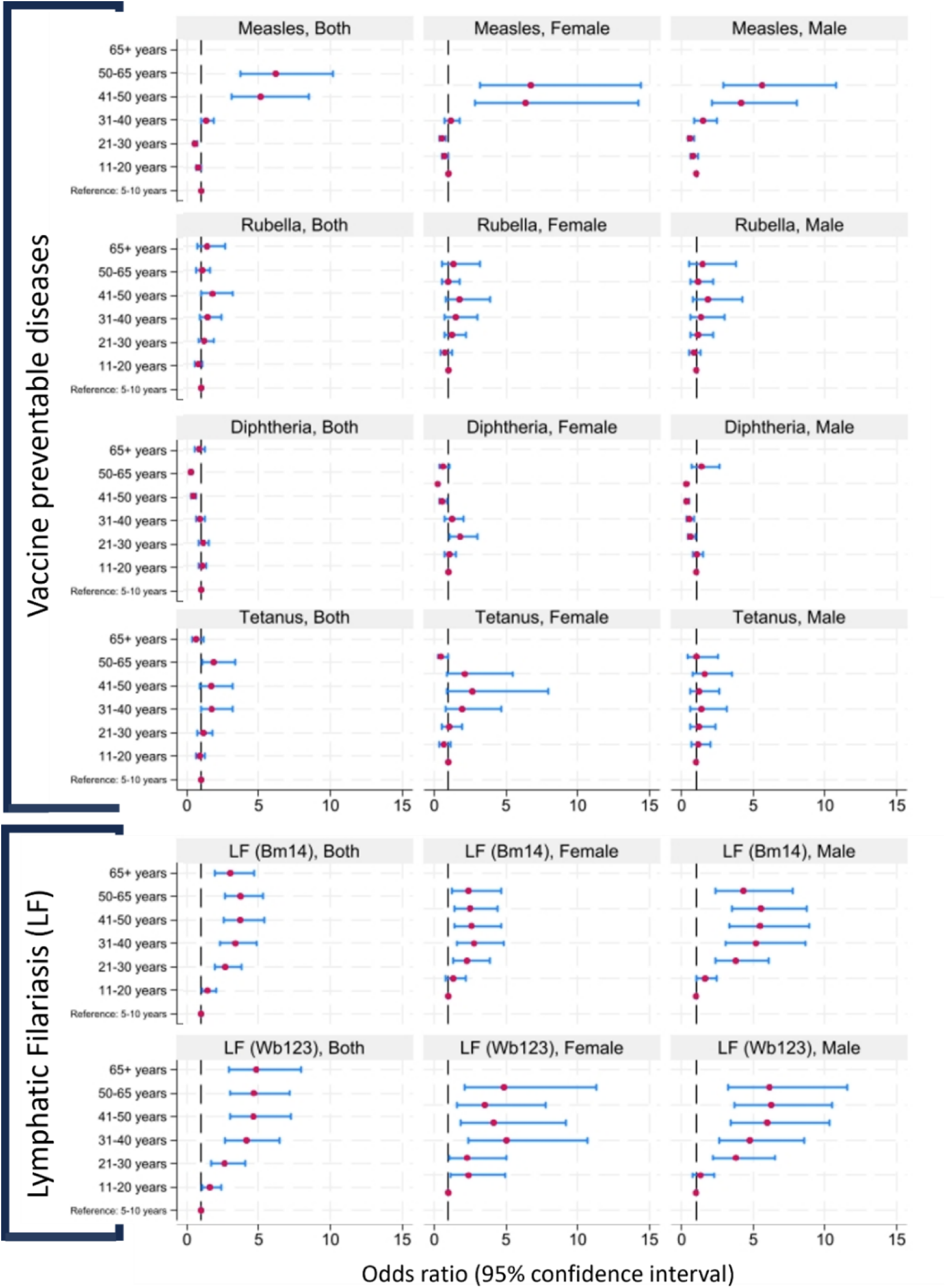
Sex and age group odds ratios for seroprevalence (reference: 5-10 years) to individual vaccine preventable diseases (measles, rubella, diphtheria and tetanus) and lymphatic filariasis (Bm14 and Wb123) by sex (female, male and both) and age group for all years combined (2018, 2019, 2023 and 2024).

#### VPDs

For all years combined, seroprevalence to measles was significantly lower in the 11-20 years (OR=0.7 [reference: 5-10 years]; 95% CI: 0.6-0.9) and 21-30 years (OR=0.5; 95% CI: 0.4-0.7) age groups, whereas seroprevalence to diphtheria was significantly lower in the 41-50 years (OR=0.9; 95% CI: 0.3-0.6) and 51-65 years (OR=0.2; 95% CI: 0.2-0.3) age groups. Regarding sex, seroprevalence was significantly higher in females for diphtheria (OR=1.5; 95% CI: 1.2-1.8).

#### NTDs

Age-group differences were not examined for trachoma (*pgp3* Ab) or yaws (*rp17* and *TmpA* Abs) as analysis was restricted to participants <14 years. For those <14 years (all years combined), no statistically significant differences in odds of seropositivity between males and females were observed. Of note, seroprevalence to *Bm14* Ab was significantly higher in males (Female OR=0.5; 95% CI: 0.4-0.6), as was seroprevalence to *Wb123* Ab (OR=0.4; 95% CI: 0.3-0.5).

## Discussion

This study examined changes in VPD and NTD seroprevalence over time in eight PSUs in Samoa, highlighting temporal trends, poly-seropositivity and age group and sex variation. Key findings include variation in VPD (measles, rubella, diphtheria, and tetanus) seroprevalence in 2023/4 compared to 2018/2019, general decline in seroprevalence to LF Abs (*Bm14* and *Wb123*) and low overall seroprevalence of trachoma (*Pgp3*) and yaws (*rp17* and *TmpA*) Abs. Upon examining polyseropositivity, this study identified only 15 individuals seronegative to all VPDs, and small number of individuals with combined seronegativity to all VPDs and seropositivity to any NTD. Statistically significant age and sex related differences were pathogen-dependant with seroprevalence to measles and rubella lowest in young adults both male and female, and older age groups for diphtheria.

Factors contributing to low seroprevalence to VPDs prior to 2019 may reflect a combination of vaccine hesitancy (24) and health system changes, whereby in 2006 healthcare was divided into the National Health Service (NHS) and MOH, resulting in fewer service provisions and reduced community engagement. In addition to the success of mass vaccination campaigns spurred by the 2019 measles outbreak (reported 134,499 MCV doses delivered between Nov 2019 and Jan 2020 (70)), the NHS and MOH were re-merged in 2019 with concentrated effort to strengthen the public health system and revitalise primary healthcare (71). The *Infants Amendment Bill* (2019) was also introduced, which outlined a legal requirement for children to be fully immunized before starting primary school (72, 73). Between 2020-2021, the COVID-19 pandemic caused large-scale disruptions to routine immunisation services (74) with an estimated 3.6 million children missing out on MCV vaccination in the broader Western Pacific region. To combat this, Samoa launched a vaccination catch-up campaign in 2023, which improved reported coverage for measles containing vaccine (MCV) at 87% for MCV-1 and 74 % for MCV-2 (75). Findings from this study reflect this increase in vaccine uptake and possible exposure to measles acquired from the 2019 outbreak. However, it is important to note that this falls short of the target to achieve and maintain a 95% population immunity in each birth cohort as set by the WHO campaign to eliminate measles in the Western Pacific region (76).

Regular examination of seroprevalence to multiple VPDs simultaneously can help gauge both national and local population immunity, detect vaccination coverage gaps or waning protection, and signal areas with emerging epidemic risk (77). Of note, the presence of discrepancies between seroprevalence to measles and rubella in 2023 may be an indication of pockets of populations that experienced higher burden of measles infection during the 2019 outbreak. The Samoan MOH Press Release in November 2019 highlighted that approximately 89% of measles cases [were] in Upolu, concentrated in districts close to the Apia Urban Area (78). Additionally, it is important to note the general decline in seroprevalence to diphtheria and age and sex group difference in seroprevalence to tetanus. Contextually, the 2019 Multiple Indicator Cluster Surveys (MICS) report highlighted a prevalence of 29.5% for children <2 years having received no Diphtheria, Tetanus and Pertussis (DTP) vaccines, of which the majority were concentrated in rural areas, compared to urban areas (79). Reported WHO/UNICEF Estimates of National Immunization Coverage (WUENIC) vaccination coverage for first dose DTP-1 is Samoa has been generally high, between 88% and 99% between the year 2010-2024, however within the same time period, DTP-3 coverage has been estimated to fluctuate between 44% and 89% (80). However, it is important to note that vaccination coverage estimates rely on self-reported history of vaccination and may under or over-estimate population immunity owing to desirability biases, inaccurate population denominators and/or administrative errors (81). This in turn highlights the value of multi-pathogen serosurveillance in complementing traditional disease surveillance programs and providing valuable information regarding public health interventions.

Currently, the WHO recommend population diphtheria-containing vaccine coverage of 80–85% to maintain community protection and reduce the threat of an outbreak (82, 83). However, immunity wanes quickly and boosters may be needed to prevent infections later in life. Regarding age group differences, the finding of low seroprevalence to diphtheria in older age groups is notable. In the context of vaccination history, older age groups would have been due to receive the initial 3 doses (≤1 year of age) of DTP vaccination between the years 1959 and 1992. It is important to note that the DTP vaccination was introduced in Samoa in 1970 (72), therefore the older portion of this age group would have likely received none, or fewer than the recommended five doses of DTP vaccine. Studies have shown that single dose efficacy only provides good protection in the first year of life, primarily owing to maternal antibodies, whereby the efficacy of two doses wanes rapidly between 2-4 years, compared to the recommended three-dose schedule and the addition of four booster doses beyond three years of age (84, 85). Furthermore, between November 2024 and May 2025, Samoa experienced an outbreak of pertussis, cumulating in over 400 cases, of which almost 40% occurred in adults aged 20+ years (86). In response, a large-scale vaccination catch up campaign was undertaken with the addition of adult DTP vaccine, Boostrix, that was previously unavailable in Samoa (87). Currently, the national immunisation policy for Samoa is approaching renewal, the inclusion of adult vaccination as a priority would strengthen alignment with the 2026 World Immunisation Weeks’s theme “For every generation, vaccines work”.

The prevalence of LF antibodies to monitor LF transmission has been previously discussed in depth in the wider Western Pacific region context (88–91), and within Samoa specifically (16, 33). Of note, the general overall decline in *Bm14* seroprevalence is reassuring given the ongoing efforts of LF elimination in Samoa (92). This pattern reflects recent trends in decline of LF prevalence from 2018 onwards; however, the persistence of pockets of increased seroprevalence may reflect communities and populations with ongoing transmission and burden (27, 31, 32). While the exact kinetics of LF Abs are not well understood, the finding of higher seroprevalence to Abs in older age groups seem to be reflective of current literature highlighting the increased burden of LF within this age group (93, 94). The utility of LF Abs as a complementary surveillance tool is gaining traction (88, 89, 95, 96), however further research is required regarding differentiation between active and past LF infection as well as seroprevalence thresholds to signal ongoing LF transmission.

Current trachoma and yaws prevalence, and the status of endemicity, is not well documented in many countries within the Pacific region (97). Historically the first documented cases of trachoma in Samoa date back to 1914, with additional research studies highlighting the gradual decline in the years of 1945 (55.1%), 1958 (5.5%), 1982 (no active cases noted), and most recently 2002 (no cases of trachoma, however 2.4% of low-vision or blindness were noted to be caused by infection or inflammation) (98–100). Until the middle of the 20th century, yaws was considered endemic in Samoa (2.9% prevalence between 1950-1960), leading to an intensive national yaws control program established in 1955 (101). In 1960, Samoa had reported 0% prevalence to yaws, while passive surveillance reported very small numbers of cases over the next few decades (101). Except for Papua New Guinea, Solomon Islands, and Vanuatu which still considered endemic, no recent new cases of yaws have been documented in the Western Pacific Region (101). However, the presence of serological antibodies warrants further attention as clinical features may identify only 30% to 60% of suspected *T. pallidum* skin ulcers (102, 103). Given that Yaws is targeted by the WHO for eradication (104), any potential cases should warrant further investigation.

A key limitation of this study is the absence of internationally accepted seropositivity thresholds for MBAs, along with the lack of formal test validation, including sensitivity and specificity, assessment of inter- and intra-assay variability, reproducibility, and consistency across batches, especially considering that 2018/2019 and 2023/2024 samples were tested in two different laboratories. This study was further limited by utilisation of samples collected for LF, and the exclusion of children aged <5 years, which may have introduced age-related bias. This, however, is a limitation of the sampling design itself, rather than a limitation of integrated serosurveillance. Additionally, surveys conducted prior to 2023 did not ask for self-reported vaccination history, which could have allowed for the comparison between seroprevalence and reported vaccination coverage. Despite these limitations, this study adds further utility of integrated serosurveillance to reveal patterns in local level seroprevalence to VPDs and NTDs.

In conclusion, this study builds upon previous work examining seroprevalence to VPDs in Samoa in 2018/2019 and highlights heterogeneous patterns of immunity to both VPDs and NTDs across PSUs, overall declining antibody responses to LF, persistently low seroprevalence to trachoma and yaws, and complex profiles of polyseropositivity, underscoring vulnerable populations with the combination of low immunity to VPDs and burden of NTDs. Despite limitations, the utilisation of MBAs for laboratory analysis enables a cost- and time-effective method for determining local-level-patterns in seropositivity to multiple VPDs and NTDs simultaneously, is ideal for use in low-resource settings, such as Samoa and other PICTs.

## Acknowledgements

We are extremely grateful to Tautala Maula and colleagues at the Samoa Red Cross for their continued and invaluable support as field teams, especially Secretary General Namulauulu Tautala Mauala, Nixon Norman Mataia, Brenda Koon Wai You, Alesi Samuelu and Shem Lepale, as well as to the staff of the Samoa Ministry of Health. We also acknowledge the significant in-country support provided by Lepaitai Hansell, Rasul Baghirov, and Dyxon Hansell at the WHO office in Apia, and extend our sincere thanks for their assistance during survey preparation and fieldwork. We sincerely thank Kimberley Won, Diana Martin, Gretchen Cooley, Ashley Simon and the teams from US CDC, for chemical coupling of beads for MBA, analysing the dried blood spot samples and calculating seropositive cut-off levels. We thank Dr Patrick Lammie and Dr Katherine Gass for their assistance in interpreting the results, and Dr Patricia Graves for her assistance with sample design and collection.

## Data availability statement

Communities in Samoa are small (some with <200 inhabitants) and sharing individual-level data that includes village and individual ages could enable identification of participants, violating the conditions of the study’s ethics approval.

## Supplementary Materials list

**Text 1:** Surveillance and Monitoring to Eliminate Lymphatic Filariasis and Scabies in Samoa (SaMELFS) study components and participant recruitment each year

**Table 1:** Seropositivity cut-off values (MFI-bg) and method of determination for antigens used in this analysis by year

**Table 2:** Calculations for population-based weighting from Samoa Census (2016 and 2021)

**Table 3:** Participant numbers by sex and age group with statistical test (chi-squared) for changes by year

**Table 4:** Odds of seroprevalence to vaccine preventable diseases (measles, rubella, diphtheria, tetanus) and neglected tropical diseases (trachoma, yaws, lymphatic filariasis) with 95% confidence interval and p-value, by year

**Table 5:** Number of participants (n; %) seropositive to various vaccine preventable disease (measles, rubella, tetanus, diphtheria) combinations by year

## References

1. Cutts FT, Hanson M. Seroepidemiology: an underused tool for designing and monitoring vaccination programmes in low-and middle-income countries. Tropical Medicine & International Health. 2016;21(9):1086–98.

2. Metcalf CJE, Farrar J, Cutts FT, Basta NE, Graham AL, Lessler J, et al. Use of serological surveys to generate key insights into the changing global landscape of infectious disease. The Lancet. 2016;388(10045):728–30.

3. Arnold BF, Scobie HM, Priest JW, Lammie PJ. Integrated serologic surveillance of population immunity and disease transmission. Emerging Infectious Diseases. 2018;24(7):1188–94.

4. Pan American Health Organization (PAHO). Toolkit for Integrated Serosurveillance of Communicable Diseases in the Americas. Washington, D.C; 2022. Contract No.: 978-92-75-12565-6.

5. Lammie PJ, Moss DM, Brook Goodhew E, Hamlin K, Krolewiecki A, West SK, Priest JW. Development of a new platform for neglected tropical disease surveillance. International Journal for Parasitology. 2012;42(9):797–800.

6. Carcelen AC, Hayford K, Moss WJ, Book C, Thuma PE, Mwansa FD, Patenaude B. How much does it cost to measure immunity? A costing analysis of a measles and rubella serosurvey in southern Zambia. PLoS ONE. 2020;15(10 October).

7. Carcelen AC, Kong AC, Takahashi S, Hegde S, Jaenisch T, Chu M, et al. Challenges and approaches to establishing multi-pathogen serosurveillance: Findings from the 2023 Serosurveillance Summit. The American Journal of Tropical Medicine and Hygiene. 2024;111(5):1145.

8. Ward S, Lawford HL, Sartorius B, Lau CL. Integrated serosurveillance of infectious diseases using multiplex bead assays: A systematic review. Tropical Medicine and Infectious Disease. 2025;10(1):19.

9. Minta AA, Andre-Alboth J, Childs L, Nace D, Rey-Benito G, Boncy J, et al. Seroprevalence of Measles, Rubella, Tetanus, and Diphtheria Antibodies among Children in Haiti, 2017. Am J Trop Med Hyg. 2020;103(4):1717–25.

10. Njenga S, Kanyi H, Mwende F, Wiegand R, Goodhew EB, Priest J, et al. Multiplex serosurveys as a tool for integrated neglected tropical diseases (NTD) surveillance. American Journal of Tropical Medicine and Hygiene. 2014;91(5):327.

11. World Health Organization. Ending the neglect to attain the Sustainable Development Goals: a road map for neglected tropical diseases 2021–2030. 2020.

12. Rathod V, Kadam L, Gautam M, Gumma PD, Marke K, Asokanathan C, et al. Multiplexed bead-based assay for the simultaneous quantification of human serum IgG antibodies to tetanus, diphtheria, pertussis toxin, filamentous hemagglutinin, and pertactin. Frontiers in Immunology. 2023;14.

13. Ward S, Lawford HL, Sartorius B, Mayfield HJ, Amosa-Lei Sam F, Sheridan SL, et al. Finding the Gaps: Integrated Serosurveillance and Spatial Clustering of Vaccine Preventable Diseases in Samoa, 2018–2019. Tropical Medicine and Infectious Disease. 2025;11(1):9.

14. Njenga SM, Kanyi HM, Arnold BF, Matendechero SH, Onsongo JK, Won KY, Priest JW. Integrated Cross-Sectional Multiplex Serosurveillance of IgG Antibody Responses to Parasitic Diseases and Vaccines in Coastal Kenya. Am J Trop Med Hyg. 2020;102(1):164–76.

15. Arzika AM, Mindo-Panusis D, Abdou A, Kadri B, Nassirou B, Maliki R, et al. Effect of Biannual Mass Azithromycin Distributions to Preschool-Aged Children on Trachoma Prevalence in Niger: A Cluster Randomized Clinical Trial. JAMA Network Open. 2022:E2228244.

16. Lawford HL, Sartorius B, Mayfield HJ, Sam FA-L, Viali S, Kamu T, et al. Sensitivity of anti-filarial antibodies for lymphatic filariasis surveillance: Insights from a serological survey in Samoa in 2018. PLOS Neglected Tropical Diseases. 2025;19(1):e0012835.

17. Organization WH. Expanded program on immunization: measles and hepatitis B. WHO/WPR/RC54/5; 2003.

18. Vynnycky E, Adams EJ, Cutts FT, Reef SE, Navar AM, Simons E, et al. Using seroprevalence and immunisation coverage data to estimate the global burden of congenital rubella syndrome, 1996-2010: a systematic review. PloS one. 2016;11(3):e0149160.

19. Group WB. Samoa Data 2025 [Available from: https://data.worldbank.org/country/samoa.

20. World Health Organization. Our work in Samoa 2025 [Available from: https://www.who.int/samoa/our-work.

21. Baghirov R, Ah-Ching J, Bollars C. Achieving UHC in Samoa through revitalizing PHC and reinvigorating the role of village women groups. Health systems & reform. 2019;5(1):78–82.

22. Sheel M, Tinessia A, Hall J, Abdi I, Craig A, Zimmerman P-A, et al. Review of the Pacific Public Health Surveillance Network, 2023. 2023.

23. Fraser-Hurt N, Naseri LT, Thomsen R, Matalavea A, Ieremia-Faasili V, Reupena MS, et al. Improving services for chronic non-communicable diseases in Samoa: an implementation research study using the care cascade framework. Australian and New Zealand Journal of Public Health. 2022;46(1):36–45.

24. Craig AT, Heywood AE, Worth H. Measles epidemic in Samoa and other Pacific islands. The Lancet Infectious Diseases. 2020;20(3):273–5.

25. Takashima Y, Aslam SK, Evans R, Mariano KM, Lee C-w, Wang X, et al. Measles and rubella elimination in the Western Pacific Region in 2013–2022: lessons learned from progress and achievements made during regional and global measles resurgences. Vaccines. 2024;12(7):817.

26. Lau CL, Meder K, Mayfield HJ, Kearns T, McPherson B, Naseri T, et al. Lymphatic filariasis epidemiology in Samoa in 2018: Geographic clustering and higher antigen prevalence in older age groups. PLoS neglected tropical diseases. 2020;14(12):e0008927.

27. Mayfield HJ, Muttucumaru R, Sartorius B, Sheridan S, Ward S, Martin BM, et al. Recurrence of microfilaraemia after triple-drug therapy for lymphatic filariasis in Samoa: recrudescence or reinfection? International Journal of Infectious Diseases. 2025.

28. Lees J, McCool J, Woodward A. Eye health outreach services in the Pacific Islands region: an updated profile. The New Zealand Medical Journal (Online). 2015;128(1420):25.

29. Butala CB, Cave RNR, Fyfe J, Coleman PG, Yang G-J, Welburn SC. Impact of COVID-19 on the neglected tropical diseases: a scoping review. Infectious Diseases of Poverty. 2024;13(1):55.

30. Graves PM, Sheridan S, Scott J, Sam FA-L, Naseri T, Thomsen R, et al. Triple-drug treatment is effective for lymphatic filariasis microfilaria clearance in Samoa. Tropical Medicine and Infectious Disease. 2021;6(2):[1].

31. Mayfield HJ, Sartorius B, Sheridan S, Howlett M, Martin BM, Thomsen R, et al. Ongoing transmission of lymphatic filariasis in Samoa 4.5 years after one round of triple-drug mass drug administration. PLOS Neglected Tropical Diseases. 2024;18(6):e0012236.

32. Mayfield HJ, Lawford H, Sartorius B, Graves PM, Sheridan S, Kearns T, et al. Epidemiology of Lymphatic Filariasis Antigen and Microfilaria in Samoa, 2019: 7–9 Months Post Triple-Drug Mass Drug Administration. Tropical Medicine and Infectious Disease. 2024;9(12):311.

33. Lawford HL, Mayfield HJ, Sam FA-L, Viali S, Kamu T, Thomsen R, Lau CL. Integrated serological surveillance for neglected tropical diseases, vaccine-preventable diseases, and arboviruses in Samoa, 2018. Scientific Reports. 2025;15(1):12667.

34. Mayfield HJ, McLure A, Rigby L, McPherson B, Bradbury R, Hedtke S, et al. Five Years, Two Mass Triple-Drug Administrations, and Ongoing Transmission: Using Mosquito and Human Indicators to Evaluate the Impact of Lymphatic Filariasis Interventions in Samoa. medRxiv. 2025:2025.12. 24.25342955.

35. Huffer E, So’o A. Beyond governance in Sâmoa: understanding Samoan political thought. The Contemporary Pacific. 2005:311–33.

36. Trade AGDoFAa. Samoa Health Design. In: Service SH, editor. Australia 2018.

37. Willis GA, Mayfield HJ, Kearns T, Naseri T, Thomsen R, Gass K, et al. A community survey of coverage and adverse events following country-wide triple-drug mass drug administration for lymphatic filariasis elimination, Samoa 2018. PLoS neglected tropical diseases. 2020;14(11):e0008854.

38. McPherson B, Mayfield HJ, McLure A, Gass K, Naseri T, Thomsen R, et al. Evaluating molecular xenomonitoring as a tool for lymphatic filariasis surveillance in Samoa, 2018–2019. Tropical medicine and infectious disease. 2022;7(8):203.

39. Chandrashekar R, Curtis K, Li B, Weil G. Molecular characterization of a Brugia malayi intermediate filament protein which is an excretory-secretory product of adult worms. Molecular and biochemical parasitology. 1995;73(1-2):231–9.

40. Weil GJ, Curtis KC, Fischer PU, Won KY, Lammie PJ, Joseph H, et al. A multicenter evaluation of a new antibody test kit for lymphatic filariasis employing recombinant Brugia malayi antigen Bm-14. Acta tropica. 2011;120:S19–S22.

41. Hamlin KL, Moss DM, Priest JW, Roberts J, Kubofcik J, Gass K, et al. Longitudinal monitoring of the development of antifilarial antibodies and acquisition of Wuchereria bancrofti in a highly endemic area of Haiti. PLoS Negl Trop Dis. 2012;6(12):e1941.

42. Pastor AF, Silva MR, Dos Santos WJT, Rego T, Brandão E, de-Melo-Neto OP, Rocha A. Recombinant antigens used as diagnostic tools for lymphatic filariasis. Parasites & Vectors. 2021;14(1):474.

43. Tisch DJ, Bockarie MJ, Dimber Z, Kiniboro B, Tarongka N, Hazlett FE, et al. Mass drug administration trial to eliminate lymphatic filariasis in Papua New Guinea: changes in microfilaremia, filarial antigen, and Bm14 antibody after cessation. The American journal of tropical medicine and hygiene. 2008;78(2):289.

44. Kubofcik J, Fink DL, Nutman TB. Identification of Wb123 as an Early and Specific Marker of Wuchereria bancrofti Infection. PLoS Neglected Tropical Diseases. 2012;6(12).

45. Steel C, Golden A, Kubofcik J, LaRue N, De los Santos T, Domingo GJ, Nutman TB. Rapid Wuchereria bancrofti-specific antigen Wb123-based IgG4 immunoassays as tools for surveillance following mass drug administration programs on lymphatic filariasis. Clinical and Vaccine Immunology. 2013;20(8):1155–61.

46. Greene SE, Huang Y, Curtis KC, King CL, Fischer PU, Weil GJ. IgG4 antibodies to the recombinant filarial antigen Wb-Bhp-1 decrease dramatically following treatment of lymphatic filariasis. PLOS Neglected Tropical Diseases. 2023;17(6):e0011364.

47. Ottesen EA, Duke B, Karam M, Behbehani K. Strategies and tools for the control/elimination of lymphatic filariasis. Bulletin of the world Health Organization. 1997;75(6):491.

48. Hussein O, Setouhy ME, Ahmed ES, Kandil AM, Ramzy RM, Helmy H, Weil GJ. Duplex Doppler sonographic assessment of the effects of diethylcarbamazine and albendazole therapy on adult filarial worms and adjacent host tissues in Bancroftian filariasis. 2004.

49. Moss DM, Priest JW, Boyd A, Weinkopff T, Kucerova Z, Beach MJ, Lammie PJ. Multiplex bead assay for serum samples from children in Haiti enrolled in a drug study for the treatment of lymphatic filariasis. Am J Trop Med Hyg. 2011;85(2):229–37.

50. Goodhew B, Tang X, Goldstein J, Lee J, Martin D, Gwyn S. Validation of immunoassays for the Chlamydia trachomatis antigen Pgp3 using a chimeric monoclonal antibody. Sci Rep. 2023;13(1):7281.

51. Goodhew EB, Priest JW, Moss DM, Zhong G, Munoz B, Mkocha H, et al. CT694 and pgp3 as serological tools for monitoring trachoma programs. PLoS Negl Trop Dis. 2012;6(11):e1873.

52. Gwyn S, Nute AW, Sata E, Tadesse Z, Chernet A, Haile M, et al. The Performance of Immunoassays to Measure Antibodies to the Chlamydia trachomatis Antigen Pgp3 in Different Epidemiological Settings for Trachoma. Am J Trop Med Hyg. 2021;105(5):1362–7.

53. Gwyn S, Mkocha H, Randall JM, Kasubi M, Martin DL. Optimization of a rapid test for antibodies to the Chlamydia trachomatis antigen Pgp3. Diagn Microbiol Infect Dis. 2019;93(4):293–8.

54. West SK, Munoz B, Kaur H, Dize L, Mkocha H, Gaydos CA, Quinn TC. Longitudinal change in the serology of antibodies to Chlamydia trachomatis pgp3 in children residing in a trachoma area. Scientific reports. 2018;8(1):3520.

55. Guagliardo SAJ, Parameswaran N, Agala N, Abubakar A, Cooley G, Ye T, et al. Treponemal Antibody Seroprevalence Using a Multiplex Bead Assay from Samples Collected during the 2018 Nigeria HIV/AIDS Indicator and Impact Survey: Searching for Yaws in Nigeria. Am J Trop Med Hyg. 2023;108(5):977–80.

56. Akins D, Purcell B, Mitra M, Norgard M, Radolf J. Lipid modification of the 17-kilodalton membrane immunogen of Treponema pallidum determines macrophage activation as well as amphiphilicity. Infection and immunity. 1993;61(4):1202–10.

57. Ijsselmuiden O, Schouls L, Stolz E, Aelbers G, Agterberg C, Top J, Van Embden J. Sensitivity and specificity of an enzyme-linked immunosorbent assay using the recombinant DNA-derived Treponema pallidum protein TmpA for serodiagnosis of syphilis and the potential use of TmpA for assessing the effect of antibiotic therapy. Journal of clinical microbiology. 1989;27(1):152–7.

58. Cooley G, Gwyn S, Priest J, Scobie H, Rogier E, Biritwum N, et al. Comparison of luminex methods for analysis of antibody responses against markers of vaccine protection and parasitic, water-borne and neglected tropical diseases. American Journal of Tropical Medicine and Hygiene. 2018;99(4):201.

59. Martin D, Cooley G, Goodhew B, Mitja O, Castro A, Chen C, et al. Development of a luminex treponemal antibody assay for large-scale surveillance and impact evaluation of global yaws eradication program. American Journal of Tropical Medicine and Hygiene. 2015;93(4):514.

60. Cooley GM, Mitja O, Goodhew B, Pillay A, Lammie PJ, Castro A, et al. Evaluation of Multiplex-Based Antibody Testing for Use in Large-Scale Surveillance for Yaws: a Comparative Study. J Clin Microbiol. 2016;54(5):1321–5.

61. Parameswaran N, Mitjà O, Bottomley C, Kwakye C, Houinei W, Pillay A, et al. Antibody responses to two recombinant treponemal antigens (rp17 and TmpA) before and after azithromycin treatment for yaws in Ghana and Papua New Guinea. Journal of clinical microbiology. 2021;59(5):10.1128/jcm.02509-20.

62. Cooley GM, Feldstein LR, Bennett SD, Estivariz CF, Weil L, Bohara R, et al. No Serological Evidence of Trachoma or Yaws Among Residents of Registered Camps and Makeshift Settlements in Cox’s Bazar, Bangladesh. Am J Trop Med Hyg. 2021;104(6):2031–7.

63. Ash VD, Hughes SJ, Liao RS. Evaluation of the Bio-Rad BioPlex 2200 MMRV multiplex flow immunoassay for the detection of IgG-Class antibodies to measles, Mumps, Rubella, and Varicella-zoster virus. Clinical Chemistry. 2011;57(10):A209–A10.

64. Coughlin MM, Matson Z, Sowers SB, Priest JW, Smits GP, van der Klis FRM, et al. Development of a Measles and Rubella Multiplex Bead Serological Assay for Assessing Population Immunity. J Clin Microbiol. 2021;59(6).

65. Lutz CS, Hasan AZ, Bolotin S, Crowcroft NS, Cutts FT, Joh E, et al. Comparison of measles IgG enzyme immunoassays (EIA)versus plaque reduction neutralization test (PRNT)for measuringmeaslesserostatus: a systematic review of head-to-head analyses of measles IgG EIA and PRNT. BMC Infectious Diseases. 2023;23(1).

66. Liang JL. Prevention of pertussis, tetanus, and diphtheria with vaccines in the United States: recommendations of the Advisory Committee on Immunization Practices (ACIP). MMWR Recommendations and reports. 2018;67.

67. Scobie HM, Khetsuriani N, Efstratiou A, Priest JW. Validation of a diphtheria toxoid multiplex bead assay for serosurveys. Diagn Microbiol Infect Dis. 2021;100(3):115371.

68. Thomas D, Dillaerts D, Cockx M, Ampofo L, She J, Desombere I, et al. Development and validation of a microfluidic multiplex immunoassay for the determination of levels and avidity of serum antibodies to tetanus, diphtheria and pertussis antigens. Journal of Immunological Methods. 2022;503.

69. Statistics SBo. 2016 CENSUS Brief No.1. In: Statistics Bo, editor. Apia, Samoa2017.

70. Organisation. UNCsFWH. Measles Outbreak in the Pacific - Situation Report No 10, January 8, 2020. Situation Report. 2020 07 Jan 2020

71. Samoa Go. Situational Analysis for the Development of the Samoa Human Resources for Health Strategy & Samoa Health Workforce Development Plan 2020/21–2025/26. In: Samoa MoH, editor. Apia, Samoa 2020.

72. Samoa Go. National Immunisation Policy 2020-2025. In: Ministry of Health, editor. Samoa 2020.

73. Thornton J. In the aftermath: the legacy of measles in Samoa. Lancet (London, England). 2020;395(10236):1535.

74. Maltezou HC, Medic S, Cassimos DC, Effraimidou E, Poland GA. Decreasing routine vaccination rates in children in the COVID-19 era. Vaccine. 2022;40(18):2525.

75. Samoa strengthens its vaccination programme [press release]. 4 July 2024 2024.

76. Organization WH. Measles vaccines: WHO position paper, April 2017–Recommendations. Vaccine. 2019;37(2):219–22.

77. Carcelen AC, Winter AK, Moss WJ, Chilumba I, Mutale I, Chongwe G, et al. Leveraging a national biorepository in Zambia to assess measles and rubella immunity gaps across age and space. Scientific reports. 2022;12(1):10217.

78. Ministry of Health Press Release 6 - Measles Epidemic [press release]. Apia, Samoa, November 22, 2019 2019.

79. Statistics SBo. Samoa demographic and health survey : multiple indicator cluster survey (DHS-MICS) 2019-20 : survey findings report. In: Division C-SaD, editor. Apia, Samoa 2021.

80. World Health Organization. Diphtheria tetanus toxoid and pertussis (DTP) vaccination coverage 2026 [Available from: https://immunizationdata.who.int/global/wiise-detail-page/diphtheria-tetanus-toxoid-and-pertussis-(dtp)-vaccination-coverage?CODE=WSM&ANTIGEN=&YEAR=.

81. Gong W, Hayford K, Taighoon Shah M, Iqbal J, Moss WJ, Moulton LH, et al. Using Serosurvey Data Triangulation for More Accurate Estimates of Vaccine Coverage: Measured and Modeled Coverage From Pakistan Household Surveys. American journal of epidemiology. 2019;188(10):1849–57.

82. Organization WH. Diphtheria vaccine: WHO position paper, August 2017–Recommendations. Vaccine. 2018;36(2):199–201.

83. Plotkin S, Robinson JM, Cunningham G, Iqbal R, Larsen S. The complexity and cost of vaccine manufacturing–an overview. Vaccine. 2017;35(33):4064–71.

84. Guiso N, Fine P. Comparative efficacy/effectiveness of schedules in infant immunisation against pertussis, diphtheria and tetanus: Systematic review and meta-analysis. 2015.

85. Vesikari T, Van Damme P, Lindblad N, Pfletschinger U, Radley D, Ryan D, et al. An open-label, randomized, multicenter study of the safety, tolerability, and immunogenicity of quadrivalent human papillomavirus (types 6/11/16/18) vaccine given concomitantly with diphtheria, tetanus, pertussis, and poliomyelitis vaccine in healthy adolescents 11 to 17 years of age. Pediatric Infectious Disease Journal. 2010;29(4):314–8.

86. Ministry of Health Pertussis Surveillance Situation Report No.8 [press release]. Apia, Samoa, 20/01/2025 2025.

87. New Zealand supports with 6000 doses of Boostrix vaccines in Samoa [press release]. Ministry of Health 2025.

88. Cadavid Restrepo AM, Gass K, Won KY, Sheel M, Robinson K, Graves PM, et al. Potential use of antibodies to provide an earlier indication of lymphatic filariasis resurgence in post–mass drug ad ministration surveillance in American Samoa. International Journal of Infectious Diseases. 2022;117:378–86.

89. Won KY, Robinson K, Hamlin KL, Tufa J, Seespesara M, Wiegand RE, et al. Comparison of antigen and antibody responses in repeat lymphatic filariasis transmission assessment surveys in American Samoa. PLoS Negl Trop Dis. 2018;12(3):e0006347.

90. Hall DD. Standardisation and commercialisation of a filarial antibody test for use in the global lymphatic filariasis elimination programme: James Cook University; 2016.

91. Yajima A, Ichimori K. Progress in the elimination of lymphatic filariasis in the Western Pacific Region: successes and challenges. International Health. 2021;13(Supplement_1):S10–S6.

92. Graves PM, Joseph H, Coutts SP, Mayfield HJ, Maiava F, Leong-Lui TAA, et al. Control and elimination of lymphatic filariasis in Oceania: Prevalence, geographical distribution, mass drug administration, and surveillance in Samoa, 1998–2017. Advances in parasitology. 2021;114:27–73.

93. Zakaria ND, Avoi R. Prevalence and risk factors for positive lymphatic filariasis antibody in Sabah, Malaysia: a cross-sectional study. Transactions of the Royal Society of Tropical Medicine and Hygiene. 2022;116(4):369–74.

94. Stolk W, Ramaiah K, Van Oortmarssen G, Das P, Habbema J, De Vlas S. Meta-analysis of age-prevalence patterns in lymphatic filariasis: no decline in microfilaraemia prevalence in older age groups as predicted by models with acquired immunity. Parasitology. 2004;129(5):605–12.

95. Lawford H, Tukia O, Takai J, Sheridan SL, Ward S, Jian H, et al. Utility of Integrated Serosurveillance for Post-Validation Surveillance of Lymphatic Filariasis: Results from a Cross-Sectional Survey in Tonga. 2025.

96. Martin BM, Brown A, Sam FA-L, Tufa A, Furuya-Kanamori L, Lau CL. The utility of Infectious Disease prevalence studies to inform public health decision-making in the Samoan Islands: A systematic review. Tropical Medicine and Infectious Disease. 2025;10(3):1–16.

97. Leckie J. Yaws and Syphilis: Forgotten Diseases of Asia-Pacific. Rockefeller Archive Center Research Report. 2019.

98. Kluxen G, Bernsmeier H. Endemic eye diseases in Samoa, 1910-1913. Klinische Monatsblatter fur Augenheilkunde. 1979;175(6):841–5.

99. Keeffe J, Taylor HR, Fotis K, Pesudovs K, Flaxman SR, Jonas JB, et al. Prevalence and causes of vision loss in Southeast Asia and Oceania: 1990-2010. British Journal of Ophthalmology. 2014;98(5):586–91.

100. Handley BL, Roberts CH, Butcher R. A systematic review of historical and contemporary evidence of trachoma endemicity in the Pacific Islands. PloS one. 2018;13(11):e0207393.

101. Capuano C, Ozaki M. Yaws in the Western pacific region: a review of the literature. Journal of tropical medicine. 2011;2011(1):642832.

102. Marks M, Chi K-H, Vahi V, Pillay A, Sokana O, Pavluck A, et al. Haemophilus ducreyi associated with skin ulcers among children, Solomon Islands. Emerging infectious diseases. 2014;20(10):1705.

103. Mitjà O, Houinei W, Moses P, Kapa A, Paru R, Hays R, et al. Mass treatment with single-dose azithromycin for yaws. New England Journal of Medicine. 2015;372(8):703–10.

104. Asiedu K, Fitzpatrick C, Jannin J. Eradication of yaws: historical efforts and achieving WHO’s 2020 target. PLoS neglected tropical diseases. 2014;8(9):e3016.

